# Generative AI and genetic analyses indicate metformin as a drug repurposing candidate for normal tension glaucoma

**DOI:** 10.1101/2024.12.02.24318301

**Authors:** Junhong Jiang, Di Hu, Qi Zhang, Zenan Lin, the μ-Biomedical Data Investigation Group (Mu-BioDig)

## Abstract

**Background:** The normal tension glaucoma (NTG) has limited drug options since current anti-glaucoma medications are mostly designed to decrease intraocular pressure (IOP). The emerging generative artificial intelligence (GAI) may provide an unprecedented approach for its drug repurposing research.

**Methods:** First, we iteratively interactivated with ChatGPT using 10 independent queries. Each query consists of two prompts, which asked ChatGPT to offer 20 drug repurposing candidates (DRCs) for NTG. The same process was employed to find DRCs with other two GAI models (i.e Google Gemini Advance and Anthropic Claude). The DRCs were quantified and ranked by their appearing frequency and orders. By tasking GAI and DrugBank database, the targets for the selected DRCs were identified. Then, the ChEMBL database was utilized to find the target-associated genes. The relevant instrumental variables (IVs) mapped to these genes were then identified with the GTEX dataset. In order to quantify the drugs’ effect, the mediation exposures (e.g. HbA1c for metformin) for the identified drugs were introduced to the single SNP mendelian randomization (SSMR) to filter the IVs with significant causal influence on the mediation traits. The filtered IVs were then utilized to measure the DRCs’ causal effect on NTG.

**Results:** Our results showed that three drugs (i.e. Metformin, Losartan, Mementine) appeared simultaneously in the suggesting lists generated by three GAI models. By utilizing GAI and DrugBank database, 8, 2 and 7 targets were identified for them, respectively. After searching ChEMBL and GTEx, the targets associated genes were identified for selecting corresponding IVs. Finaly, the SSMR kept 308 IVs for metformin, 11 for losartan, 180 for memantine. Applying the target-based MR, we found that, metformin may exert causal influence on NTG through targets GLP-1 and gluconeogenic enzymes, while no obvious causal links were detected in the study on losartan and mementine.

**Conclusions:** Our results offered novel evidences to support the metformin’s repurposing in NTG patients. Moreover, we firstly proposed a novel paradigm consisting of GAI and genetic tools, which could serve as an effective pipeline for drug repurposing investigations of other diseases.

## 1. Introduction

Glaucoma is a group of optic neuropathies characterized by progressive degeneration of retinal ganglion cells and irreversible loss of the visual field.^1^ As the primary cause of irreversible blindness worldwide, glaucoma affects more than 70 million people worldwide, with approximately 10% being bilaterally blind, imposing a significant economic burden on individuals and healthcare systems.^2, 3^ Elevated intraocular pressure (IOP) is well recognized as the leading risk factor that contributes to glaucoma development and progression, and clinically, current treatment strategies for glaucoma mainly focus on pharmacological, laser, or surgical therapies to lower IOP.^4^ However, a significant proportion of primary open-angle glaucoma patients, referred to as normal-tension glaucoma (NTG), still present progressive optic neuropathies when their IOP levels fluctuated within the normal range, and IOP reduction alone is not sufficient to halt or delay progression in all individuals with NTG.^5^ Although the role of IOP-independent risk factors in NTG pathogenesis has been recognized, there is still a lack of safe and effective therapy for NTG, except for IOP reduction in the clinic. Thus, there is an urgent need to develop novel alternative therapeutic strategies for NTG.

Drug repurposing, which refers to evaluating approved or investigational drugs for their possible new medical indications outside the original scope, significantly shortening the drug development timelines, reducing overall development costs, and avoiding risks compared to traditional drug discovery, is receiving increased interest as an alternative drug discovery strategy and has been successful for cancer, multiple sclerosis, and HIV treatment.^6^ Identifying drug repurposing candidates (DRCs) is labor-intensive and time-consuming. Based on a comprehensive review of the scientific literature concerning the safety and efficacy of drugs from the perspectives of disease mechanisms, molecular biology, pharmacology, and so on.^7^

Technological advances in artificial intelligence (AI) in recent years have begun to transform the fields of biology and medicine.^8^ Generative AI, such as ChatGPT, a class of large language models based on machine learning algorithms and natural language processing, enables large-scale capturing, processing, and analyzing various information from multi-dimensional datasets and then understanding and responding to diverse inquiries.^9^ Now, generative AI has started to play an increasingly vital role in healthcare and medicine, including protein structure prediction and novel drug discovery.^10^ Drug target-based Mendelian randomization, a powerful statistical method that uses the significantly associated single nucleotide polymorphisms as instrumental variables, has been used to find the drug-repurposing candidates for various diseases.^11^ This study aims to utilize generative AI to screen for potential DRCs for NTG and to validate these candidates using drug target-based Mendelian randomization analyses.

## 2. Methods

### 2.1 Interaction with GAI models for drug repurposing candidates (DRCs)

In a recent study^12^ on finding DRCs, Yan and colleagues proposed an effective and succinct method to summarize the findings. They interacted with ChatGPT-4 with 10 independent queries. Each query was composed of two prompts, of which the first described the details for generating drug repurposing candidates, the second asked ChatGPT to refine its previous result, and to regenerate a new output when necessary. This two-step strategy was employed in our study. Though Yan et al. only used ChatGPT and got fine results, we supposed that a combination of the findings from other GAI models would provide a more comprehensive landscape. Thus, we performed the same analysis with Anthropic Claude 3.5 and Google Gemini Advance. All queries were conducted from inception to 2024.07.13. The query was shown as follows. The screenshot of the example query can be seen in supplementary Figure S1.

- **Prompt 1:** Please offer a list of 20 most promising drug candidates for repurposing in treating normal tension glaucoma based on their potential efficacy, and show the original diseases they were designed to treat. Please demonstrate the findings in a descending rank order according to their potential effectiveness in a JSON format containing the drugs’ names and corresponding diseases.
- **Prompt 2:** Please make sure that the results meet the following inclusion criteria: 1. the drugs that were originally developed for any kinds of glaucoma should be excluded; 2. the drugs were distinct; 3. they were demonstrated in a descending order based on the potential efficacy. If the list items were less than 20, please reproduce a list of 20 valid drugs. The detailed cutoff date of the AI model should be remarked at the end of the list.

The study with ChatGPT4o offered 10 lists of DRCs, of which each list consisted of 20 candidates. The DRCs were integrated, quantified and ranked by their appearing frequency and orders. Then, a final list of unique DRCs was generated. Similarly, Anthropic Claude 3.5 and Google Gemini Advance also offered a list of ordered DRCs. In the end, we selected those DRCs which were identified simultaneously in the list of three GAI models.(See supplementary Table S1-S3)

### 2.2 Identification of the DRCs-associated instrumental variables (IVs)

The abovementioned process provided three drug candidates (Metformin, Losartan and mementine). Then, we interacted with ChatGPT with the following simple prompt.

**Prompt 1:** Can you help list the drug targets for Drug example (i.e. Metformin, Losartan and Mementine)?

Afterwards, we searched the DrugBank database (https://go.drugbank.com/drugs)^13^ to gain the drug targets for these three drugs. The target results generated from ChatGPT and DrugBank were aggregated.(See supplementary Table S4-S6) Finally, the ChEMBL (https://www.ebi.ac.uk/ChEMBL/)^14^ and the Genotype-Tissue Expression (GTEx)^15^ project were utilized to find target-associated genes (See supplementary Table S7-S9) and their relevant IVs, respectively.(See supplementary Table S10-S17 for metformin, Table S18-S19 for Losartan, Table S20-Table S26 for Mementine)

### 2.3 Selection of the IVs

Three identified drugs had their primary indications and acknowledged effects, i.e. HbA1c for metformin, blood pressure for losartan and cognitive performance for memantine. In order to quantify the drugs’ effect of each drug, the MR studies were employed to filter the DRCs-associated IVs which had significant causal influence on HbA1c, systolic blood pressure and cognitive performance. Their GWAS summary statistics were accessed from the IEU GWAS platform (GWAS ID: ebi-a-GCST90014006, ieu-b-38, Davies2018_UKB_RT_summary_results_29052018.txt respectively.) The detailed information of these traits were summarized on supplementary Table S27.

### 2.4 The outcome dataset of normal tension glaucoma NTG

The GWAS datasets for NTG were acquired from the Finnish biobank project FinnGen (https://www.finngen.fi/fi). The study included 432845 participants of European descent, with 2595 being NTG patients and 430250 being healthy controls. Their GWAS summary statistics data were acquired through the platform of the FinnGen (https://storage.googleapis.com/finngen-public-data-r11/summary_stats/finngen_R11_H7_GL AUCOMA_NTG.gz, ID: “H7_GLAUCOMA_NTG”)^16^. It was publicly released on June 24, 2024.

### 2.5 MR analysis and meta-analysis

In the MR analysis, the inverse-variance weighted (when IVs>2) and Wald ratio (when IVs≤2) approaches were selected to be the major analytical tools. Other sensitivity methods such as MR Egger, Weighted median, Weighted mode and Simple mode were employed to assess the robustness of the conclusions of the MR analysis with IVs>2. The fixed-effect and random-effect statistical models were employed for the meta-analysis. All analyses were conducted with R packages ieugwasr (version 0.1.5), TwoSampleMR (version 0.5.6), and meta (version 6.0-0). A *P* value less than 0.05 was considered statistically significant. Meta-analysis was performed with the fixed- and random-effect models.

## 3. Results

### 3.1 The identification of DRCs for NTG

As shown in Figure 1., 10 queries were performed independently in three GAIs (ChatGPT, Google Gemini Advance, Anthropic Claude 3.5). Each query generated 20 Drugs. They were assigned a score (From 1 to 20) according to their orders. After 10 independent query, the appearing frequency and average score of each drug were calculated. Those with the high frequency (maximum 10) and minimal average score were identified to be the most potential repurposing candidates. These analytical processes were conducted with ChatGPT, Anthropic Claude and Google Gemini Advance simultaneously (See supplementary Figure S1 and Table S1-S3). In the end, only three drugs (Metformin, Losartan and Mementine) appeared in the results of three GAI models.

**Figure 1.**
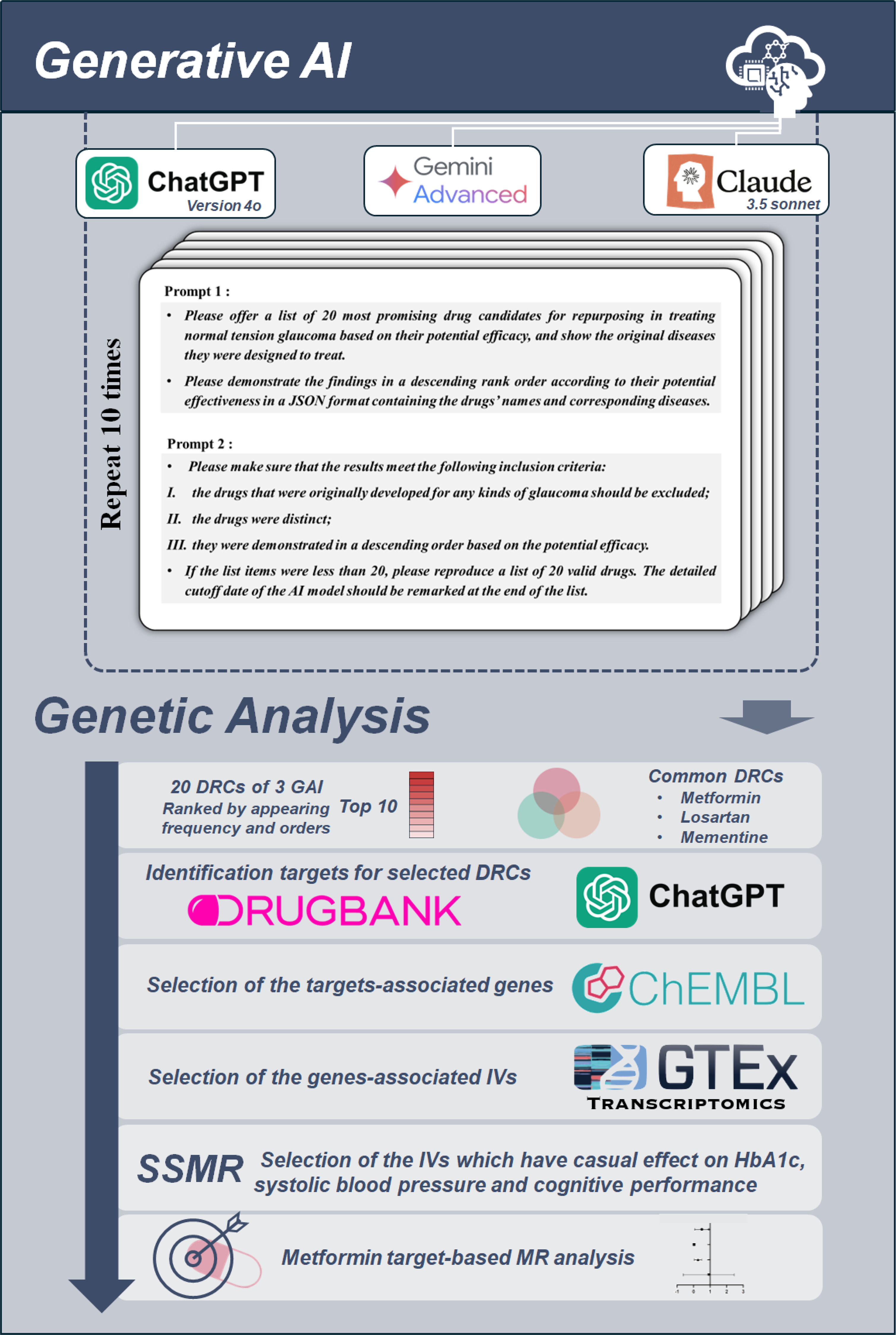
The workflow for the current study.

### 3.2 Identification of the IVs for Metformin, Losartan and Mementine

By using GAI and DrugBank, 8, 2 and 7 targets were identified for the metformin, losartan and memantine, respectively. (See supplementary Table S10-S26)

After searching ChEMBL, we found 70 target-associated genes for metformin. (See supplementary Table S28) Losartan and memantine had 2 (*AGTR1* and *AGTR2*) and 74 target-associated genes, respectively. (See supplementary Table S29) Then, the GTEx database was introduced to find IVs for each gene. (See supplementary Table S10-S26)

### 3.3 Optimizing the IVs by single SNP mendelian randomization (SSMR)

Previous studies had confirmed the Glucose-lowering influence of metformin. Thus, HbA1c was selected as the mediation trait to filter those IVs with glucose-lowering effect. After SMMR, 308 IVs with significant causal effect on HbA1c level were kept for further analysis on metformin. (see supplementary Table S30.) Similarly, the mediation trait systolic blood pressure and cognitive performance were selected for the analyses of Losartan and Mementine. In the end, 11 and 180 IVs remained for Losartan and Mementine. (See supplementary Table S31-S32)

### 3.4 Metformin functions to target NTG through targets GLP-1 and Gluconeogenic enzymes

By utilizing drug target-based MR, no obvious causal influences were found between the losartan- and mementine-associated targets and NTG.(See supplementary Table S33 and Table S34) However, the glucose-lowering effect of metformin were identified to have causal effect on NTG through two targets Gluconeogenic enzymes (Inverse variance weighted method, Beta value=-4.8075, *P*=4.3110E-05) and GLP-1 (Inverse variance weighted method, Beta value=-0.9839, *P*=0.0465). (See Fig 2.A and B and supplementary Table S35) In the meta-analysis, though the random effect model indicated no significant influence of metformin targets on reducing NTG’s risk, the common effect model suggested a remarkable effect of metformin (through GLP-1 and Gluconeogenic enzymes) on decreasing NTG’s risk (see Fig. 2C).

**Figure 2.**
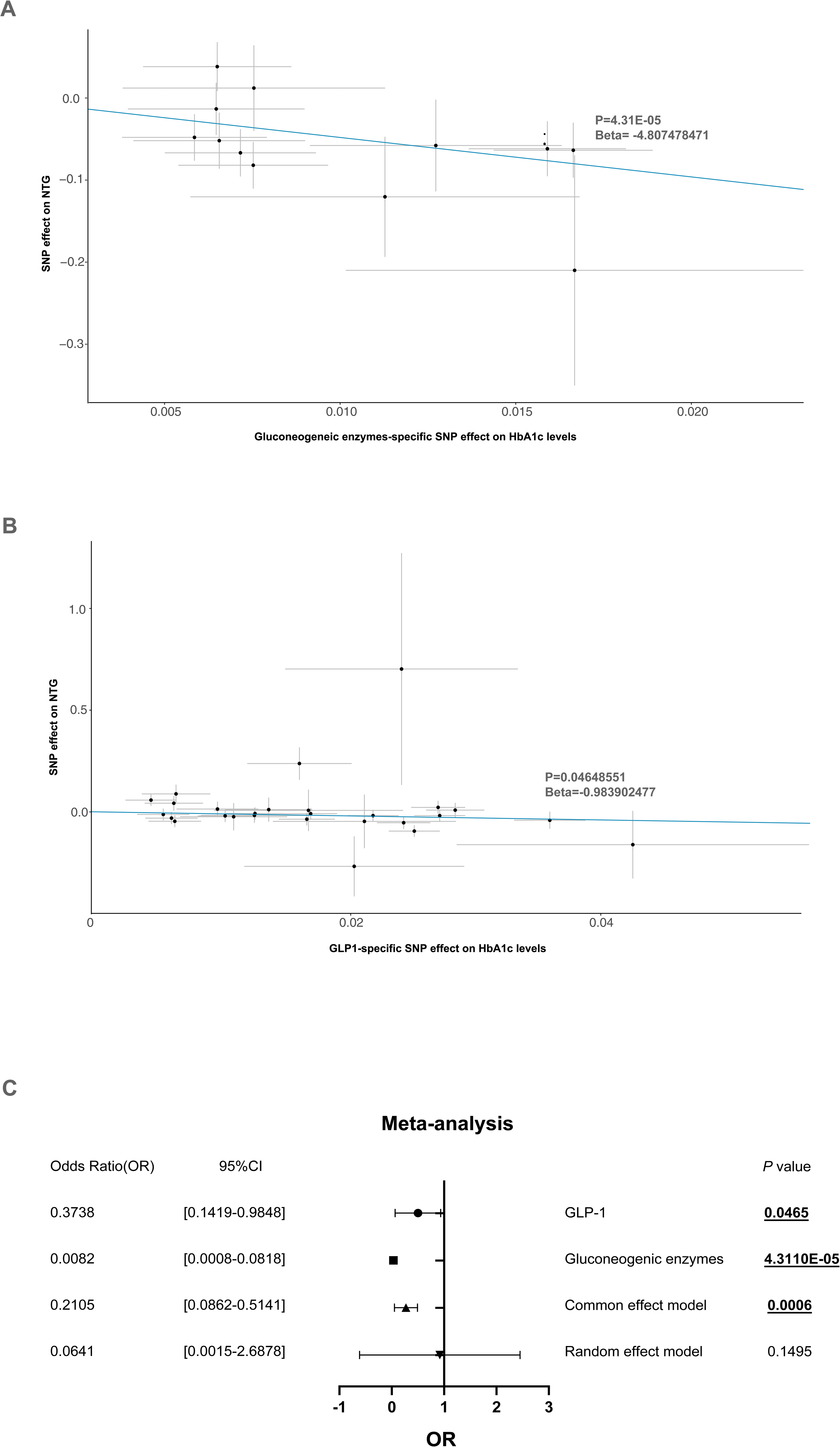
The MR analysis of Gluconeogenic enzymes- and GLP-1-specific metformin’s effect on NTG (A and B). The meta-analysis of two targets-specific metformin’s effect on NTG (C).

## 4. Discussion

Since its first reported use in the 1850s,^17^ the NTG treatment remains a dilemma for both patients and physicians. This study demonstrates for the first time the potential use of metformin as a drug repurposing candidate for NTG using a comprehensive screening strategy based on generative AI and genetic analyses. Our analyses provide a novel insight into potential targets for repurposing or de novo drug development for NTG therapies.

Metformin, the first-line treatment for type 2 diabetes, reduces blood glucose levels by multiple mechanisms. As the role of metformin in anti-inflammatory, neuromodulation, vasoregulation, and reducing oxidative stress are revealed, metformin has been gradually used in the treatment of cancer, aging, and cardiovascular disease in recent years.^18^ Notably, vasoregulation^19^ and oxidative stress^20^ are also the major leading factors for the pathogenesis of NTG. Large cohort studies and meta-analyses have shown that hyperglycemia is consistently associated with higher IOP and a significantly increased risk of glaucoma.^21, 22^ Experimental results revealed that hyperglycemia may lead to retinal ganglion cell endoplasmic reticulum stress in glaucoma.^23^ Accumulating evidence suggests that metformin interventions may be beneficial in delaying glaucoma progress. A 10-year large retrospective study from the United States managed care network comprising 150016 patients showed that the use of metformin in patients with diabetes mellitus could reduce the risk of open-angle glaucoma development.^24^ Similarly, the Rotterdam study, a prospective, population-based study that included 11260 participants, showed that patients with type 2 diabetes treated with metformin were associated with a lower risk of open-angle glaucoma.^25^ However, there has been a paucity of information regarding whether the treatment of metformin benefits NTG. Metformin could lower glucose by suppressing gluconeogenic enzymes and increasing glucagon-like-peptide-1 (GLP-1).^26^ The results of this study showed that metformin may also deliver therapeutic effects on NTG through glucagon and GLP-1, which are the two mediators.

GLP-1 is an incretin hormone secreted by intestinal L-cells that acts on the GLP-1 receptor (GLP-1R) to lower blood glucose by inhibiting glucagon secretion and enhancing insulin production and secretion.^27^ (GLP-1R) is widely expressed in various cells and tissues, including kidney, lung, heart, endothelial cells, neurons, and astrocytes, as well as immune cells. Recently, growing research demonstrated that GLP-1 receptor agonists have been shown to improve cardiovascular outcomes and attenuate atherosclerotic lesions via reducing vascular endothelial dysfunction and inhibiting atherosclerosis and inflammation.^28, 29^ Furthermore, GLP-1 receptor agonists have also been shown to protect against Parkinson’s and Alzheimer’s disease by enhancing neurotransmission and inhibiting neuroinflammation.^30^ More recently, a nationwide cohort study in Denmark found evidence of a reduction in the development of glaucoma among individuals with type 2 diabetes exposed to GLP-1 receptor agonists treatment.^31^ A similar protective effect of GLP-1 receptor agonists on glaucoma has been observed in a retrospective study covering a period of 18 years in the United States.^32^ The above researches indirectly support the findings of the present study that metformin may act as a potential therapeutic candidate for NTG. Further prospective discovery work and validation studies are needed to validate.

Our findings suggest that GAI and genetic tools could serve as an effective pipeline for drug repurposing. The powerful large-scale data mining and processing of GAI can effectively improve drug screening, and has been successfully employed in the initial screening of drug developments for Alzheimer’s disease.^10^ Yan et al. made use of ChatGPT to identify drug repurposing candidates for Alzheimer’s disease and further validated using real-world clinical data;^10^ this new mode provides a cost-effective pipeline to investigate preliminary drug screening. Drug target-based Mendelian randomization, a powerful statistical method that uses the significantly associated single nucleotide polymorphisms as instrumental variables, has been used to find drug-repurposing candidates for various diseases.^11^ In previous research, our research group screened out the potential drug for hypertension through a drug target-based Mendelian randomization analysis.^33^ In this study, we combine GAI preliminary drug screening with drug target-based Mendelian Randomization analysis to bring forward a novel approach for drug repurposing investigations.

Some strengths and limitations of the present study that deserve mentioning. First, this study used multiple mainstream GAI models to identify the drug repurposing candidates for NTG, making the screening results more comprehensive and systematic. Second, this study validates the effect of metformin treatment by estimating the causal effect of metformin on NTG using drug target-based MR, which could reduce biases and confounding seen in non-genetic observational studies. However, our study analyzed only the data from European participants, and these genetic analyses are not a substitute for the conducted randomized clinical trial. Further confirmation from prospective studies is still in urgent need.

## 5. Conclusions

In summary, the present study provided novel evidences to support the metformin’s repurposing in NTG patients. We revealed that metformin might decrease NTG risk through lowering glucose via GLP-1 and gluconeogenic enzymes. Furthermore, we first proposed a novel paradigm consisting of GAI and genetic tools, which could serve as an effective pipeline for preliminary drug repurposing investigations before initiating clinical development.

## Data Availability

The links of the GWAS data were described appropriately in the paper. The codes and detailed information required to replicate the results in this work are available from the corresponding authors upon reasonable request.

## Author Contributions

Conception, supervision and administration: Zenan Lin and Qi Zhang; Data curation: Junhong Jiang and Di Hu; Investigation: Junhong Jiang and Di Hu; Methodology: Zenan Lin and Qi Zhang; Writing – original draft: Junhong Jiang and Di Hu; Writing – review & editing: Zenan Lin and Qi Zhang.

## Ethics Approval

The GWAS data were publicly available and approved by their original institutions. An ethics approval for the current work is not required.

## Acknowledgement

We want to acknowledge the participants and investigators of the UK Biobank, FinnGen, GTEx who made the GWAS summary statistics publicly available. We also want to thank the researchers of and DrugBank and ChEMBL who made valuable scientific data publicly available.

