## Supplementary Figure S1, Table S1-S35 for "Generative AI and genetic analyses indicate metformin as a drug repurposing candidate for normal tension glaucoma": Figure S1.pdf

ChatGPT

Please offer a list of 20 most promising drug candidates for repurposing in treating normal tension glaucoma based on their potential efficacy, and show the original diseases they were designed to treat. Please demonstrate the findings in a descending rank order according to their potential effectiveness in a JSON format containing the drugs' names and corresponding diseases.

Searched 6 sites

Based on recent research and clinical trials, here is a list of the 20 most promising drug candidates for repurposing in treating normal tension glaucoma (NTG), ranked by their potential efficacy. The original diseases they were designed to treat are also included.

json Copy code


```json
[
  {
    "drug_name": "Donepezil",
    "original_disease": "Alzheimer's Disease"
  },
  {
    "drug_name": "Bimatoprost",
    "original_disease": "Glaucoma (general)"
  },
  {
    "drug_name": "Citicoline",
    "original_disease": "Stroke Recovery"
  },
  {
    "drug_name": "Memantine",
    "original_disease": "Alzheimer's Disease"
  },
  {
    "drug_name": "Brimonidine",
    "original_disease": "Ocular Hypertension"
  },
  {
    "drug_name": "Latanoprost",
    "original_disease": "Glaucoma (general)"
  },
  {
    "drug_name": "Travoprost",
    "original_disease": "Ocular Hypertension"
  },
  {
    "drug_name": "CoQ10 (Coenzyme Q10)",
    "original_disease": "Heart Conditions"
  },
  {
    "drug_name": "Riluzole",
    "original_disease": "Amyotrophic Lateral Sclerosis (ALS)"
  },
  {
    "drug_name": "Metformin",
    "original_disease": "Type 2 Diabetes"
  },
  {
    "drug_name": "Nicotinamide",
    "original_disease": "Vitamin B3 Deficiency"
  },
  {
    "drug_name": "Pyruvate",
    "original_disease": "Metabolic Disorders"
  },
  {
    "drug_name": "Durysta (Bimatoprost Implant)",
    "original_disease": "Glaucoma (general)"
  },
  {
    "drug_name": "NAD+ Precursors",
    "original_disease": "Mitochondrial Dysfunction"
  },
  {
    "drug_name": "Melatonin",
    "original_disease": "Sleep Disorders"
  },
  {
    "drug_name": "Omega-3 Fatty Acids",
    "original_disease": "Cardiovascular Disease"
  },
  {
    "drug_name": "Resveratrol",
    "original_disease": "Aging and Chronic Diseases"
  },
  {
    "drug_name": "Cilostazol",
    "original_disease": "Peripheral Artery Disease"
  },
  {
    "drug_name": "Minocycline",
    "original_disease": "Bacterial Infections"
  },
  {
    "drug_name": "Erythropoietin",
    "original_disease": "Anemia"
  }
]
```


These drug candidates have shown potential in neuroprotection and reducing intraocular pressure, key factors in managing NTG. Donepezil, an Alzheimer's drug, has demonstrated significant neuroprotective effects on retinal ganglion cells, which are crucial for NTG treatment PENTAVISION REVIEW OF OPTOMETRY . Similarly, drugs like citicoline and memantine, originally used for stroke recovery and Alzheimer's respectively, have shown promise in protecting optic nerve cells and improving mitochondrial function AMERICAN ACADEMY OF OPHTHALMOLOGY REVIEW OF OPTOMETRY .

Please make sure that the results meet the following inclusion criteria: 1. the drugs that were originally developed for any kinds of glaucoma should be excluded; 2. the drugs were distinct; 3. they were demonstrated in a descending order based on the potential efficacy. If the list items were less than 20, please reproduce a list of 20 valid drugs. The detailed cutoff date of the AI model should be remarked at the end of the list.

Based on the updated inclusion criteria, here is a list of 20 distinct drug candidates for repurposing in treating normal tension glaucoma (NTG), excluding those originally developed for any types of glaucoma, ranked by their potential efficacy:

json Copy code


```json
[
  {
    "drug_name": "Donepezil",
    "original_disease": "Alzheimer's Disease"
  },
  {
    "drug_name": "Memantine",
    "original_disease": "Alzheimer's Disease"
  },
  {
    "drug_name": "Citicoline",
    "original_disease": "Stroke Recovery"
  },
  {
    "drug_name": "CoQ10 (Coenzyme Q10)",
    "original_disease": "Heart Conditions"
  },
  {
    "drug_name": "Riluzole",
    "original_disease": "Amyotrophic Lateral Sclerosis (ALS)"
  },
  {
    "drug_name": "Metformin",
    "original_disease": "Type 2 Diabetes"
  },
  {
    "drug_name": "Nicotinamide",
    "original_disease": "Vitamin B3 Deficiency"
  },
  {
    "drug_name": "Pyruvate",
    "original_disease": "Metabolic Disorders"
  },
  {
    "drug_name": "NAD+ Precursors",
    "original_disease": "Mitochondrial Dysfunction"
  },
  {
    "drug_name": "Melatonin",
    "original_disease": "Sleep Disorders"
  },
  {
    "drug_name": "Omega-3 Fatty Acids",
    "original_disease": "Cardiovascular Disease"
  },
  {
    "drug_name": "Resveratrol",
    "original_disease": "Aging and Chronic Diseases"
  },
  {
    "drug_name": "Cilostazol",
    "original_disease": "Peripheral Artery Disease"
  },
  {
    "drug_name": "Minocycline",
    "original_disease": "Bacterial Infections"
  },
  {
    "drug_name": "Erythropoietin",
    "original_disease": "Anemia"
  },
  {
    "drug_name": "Ibuprofen",
    "original_disease": "Inflammation/Pain"
  },
  {
    "drug_name": "Simvastatin",
    "original_disease": "Hypercholesterolemia"
  },
  {
    "drug_name": "Acetyl-L-carnitine",
    "original_disease": "Mitochondrial Dysfunction"
  },
  {
    "drug_name": "Ginkgo Biloba",
    "original_disease": "Cognitive Disorders"
  },
  {
    "drug_name": "Tamoxifen",
    "original_disease": "Breast Cancer"
  }
]
```


This list adheres to the criteria by excluding drugs originally developed for glaucoma, ensuring each drug is distinct, and ordering them based on potential efficacy as inferred from the sources reviewed.

**Cutoff date of AI model: July 2023**

Message ChatGPT

ChatGPT can make mistakes. Check important info.
